# Definition of normal vertebral morphology using NHANES-II radiographs

**DOI:** 10.1101/2022.05.05.22274448

**Authors:** John A. Hipp, Trevor F. Grieco, Patrick Newman, Charles A. Reitman

**Author notes:** Corresponding Author: John A. Hipp, PhD, Email address, Mailing address: 1632 Summit Lake Shore Rd, NW, Olympia, WA 98502.

## Abstract

**Background:** A robust definition of normal is required to confidently identify vertebral abnormalities such as fractures. Between 1976 and 1980, the 2nd National Health and Nutrition Examination Survey (NHANES-II) was conducted. Justified by the prevalence of neck and back pain, approximately 10,000 lateral cervical spine and 7,000 lateral lumbar spine X-rays were collected. Demographic, anthropometric, health, and medical history data were also collected. This resource can be used for establishing normative reference data that can subsequently be used to diagnose abnormal vertebral morphology.

**Purpose:** 1) Develop normative reference data for vertebral morphology using the lateral spine radiographs from NHANES-II. 2) Document sources of variability.

**Subject Sample:** Nationwide probability sample to document health status of the United States.

**Methods:** The coordinates of the four vertebral body corners were obtained using previously validated, automated technology consisting of a proprietary pipeline of neural networks and coded logic. These landmarks were used to calculate six vertebral body morphology metrics: 1) anterior/posterior vertebral body height ratio (VBHR); 2) superior/inferior endplate width ratio (EPWR); 3) forward/backward diagonal ratio (FBDR); 4) height/width ratio (HWR); 5: angle between endplates (EPA); 6) Angle between posterior wall and superior endplate (PSA). Descriptive statistics were generated and used to identify and trim outliers from the data and obtain a gaussian distribution for each metric. Descriptive statistics were tabulated using the trimmed data for use in quantifying deviation from average for each metric. The dependency of these metrics on sex, age, race, nation of origin, height, weight, and BMI was also assessed.

**Results:** Computer generated lumbar landmarks were obtained for 42,980 vertebrae from lumbar radiographs and 54,093 vertebrae from cervical radiographs for subjects 25 to 74 years old. After removing outliers, means and standard deviations for the remaining 35,275 lumbar and 44,938 cervical vertebrae changed only slightly, suggesting that normal morphology and intervertebral alignment is dominant in the data. There was low variation in vertebral morphology after accounting for vertebra (L1, L2, etc.), and the R^2^ was high for analyses of variance. The EPWR, FBDR and PSA generally had the lowest coefficients of variation. Excluding outliers, Age, sex, race, nation of origin, height, weight, and BMI were statistically significant for most of the variables, though the F-statistic was very small compared to that for vertebral level. Excluding all variables except vertebra changed the R^2^ very little (e.g. for the lumbar data, VBHR R^2^ went from 0.804 to 0.795 and FBDR R^2^ went from 0.9005 to 0.9000). Reference data were generated that can be used to produce standardized metrics in units of standard deviation from average. This allows for easy identification of abnormalities resulting from vertebral fractures, atypical vertebral body morphologies, and other congenital or degenerative conditions. Standardized metrics also remove the effect of vertebra thereby enabling data for all vertebrae to be pooled in research studies.

**Conclusions:** The NHANES-II collection of spine radiographs and associated data may prove to be a valuable resource that can facilitate standardized spine metrics useful for objectively identifying abnormalities. The data may be particularly valuable for identification of vertebral fractures, although X-rays taken early in life would be needed in some cases to differentiate between normal anatomic variants, fractures, and vertebral shape remodeling.

## Introduction

Although vertebral fractures can be asymptomatic, these fractures can be a source of symptoms. One of the most important predictors of subsequent fractures is the number of prior fractures.^(1-3)^ Vertebral fractures are therefore diagnostically important. Fractures can be missed in clinical practice^(4,5)^. Ferrar et al. succinctly identified the challenge of fracture detection: “The identification of vertebral fractures is problematic because (1) ‘‘normal’’ radiological appearances in the spine vary greatly both among and within individuals; (2) ‘‘normal’’ vertebrae may exhibit misleading radiological appearances due to radiographic projection error; and (3) ‘‘abnormal’’ appearances due to non-fracture deformities and normal variants are common, but can be difficult to differentiate from true vertebral fracture.”^(6)^

A typical definition of a fracture is “20% or greater loss in expected vertebral body height.” This definition requires definition of “expected” vertebral body height and that requires accurate measurements of vertebral body shape in a large and representative population. Validation of the clinically meaningful threshold level for change in vertebral dimensions also requires a reliable reference standard. Inaccuracies in measuring shape and the challenge of finding a suitable reference standard are well documented. ^(6-43)^ One option is to use the patient’s own adjacent vertebral bodies as a reference, but there is no definitive method to assure that the adjacent levels are normal, or reference data to know how much variability between levels is normal. Normative vertebral body shape metrics that represent a large proportion of the population and account for age, sex, and other confounding factors have not been previously published. Robust reference data and an understanding of the important confounding factors would facilitate more accurate and reproducible diagnosis of the presence and severity of vertebral body fractures.

Between 1976 and 1980, the 2nd National Health and Nutrition Examination Survey (NHANES-II) was conducted^1^. This was a nationwide probability sample to document health status of the United States. Justified by the prevalence and societal impact of neck and back pain, approximately 10,000 lateral cervical spine and 7,000 lateral lumbar spine X-rays were collected as part of this survey. Demographic, anthropometric, health, and medical history data were also collected. This resource can be used for establishing normative reference data that can subsequently be used to diagnose abnormalities such as fractures, vertebral deformities, and congenital abnormalities. These data can also be used to determine if the sex, age, race, height, weight and/or body mass index (BMI) of an individual are needed to predict normal vertebral morphology.

## Methods

The NHANES-II images and data were obtained through public access^2^. A previously validated series of neural networks and coded logic (Spine Camp™, Medical Metrics, Inc., Houston, TX) were used to obtain four landmarks for each vertebral body from L1 to S1 (Figure 1). Although the NHANES-II lumbar X-rays almost always included up to T10 in the field-of-view, the neural networks were only trained to recognize L1 to S1. The neural networks include quality control networks to assure the X-ray was a lateral lumbar view, manipulate the image if needed so upper vertebrae are toward the top of the X-ray, with spinous processes toward the left side, and with bone being whiter than air. Neural networks and coded logic were then used to segment the bone, find individual vertebrae, label the vertebrae, find the four corners, and finally refrain from reporting landmark coordinates when the confidence of the networks was too low. All the neural networks were trained with over 50 thousand lateral X-rays where analysts had previously digitized standardized landmarks. The NHANES images were not used in training the neural networks. The analysts used Quantitative Motion Analysis software (QMA^®^, Medical Metrics, Inc., Houston, TX) to place the landmarks. The QMA^®^ software has previously been validated ^(44-49)^.

**Figure 1:**
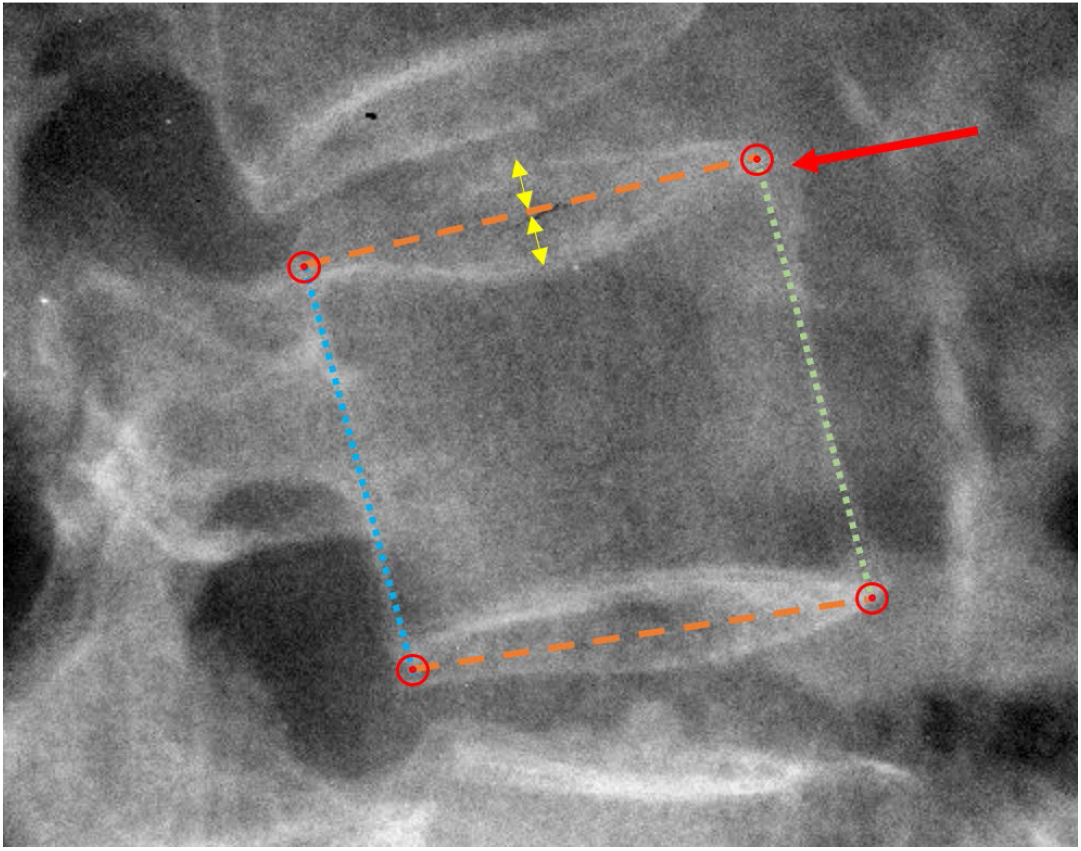
Details of anatomic landmark placement. Dashed lines show the assumed mid-sagittal plane of the superior and inferior endplates, identified as bisecting the radiographic shadows of the left and right sides of the endplates (yellow arrows). The red circles show the four landmarks used to measure vertebral body morphology. The red arrow points to an anterior osteophyte that is ignored. The dotted lines show the anterior and posterior vertebral body heights.

Details of landmark placement are provided, since as Keynan et al. noted, “There is a lack of standardization in the literature regarding choice and technique for the measurement of these parameters.”^(37)^. The standardized landmark placement was focused on the mid-sagittal plane of each vertebral body (Figure 1), excluding residual uncinate processes, and placing landmarks that identify the corners of the vertebral bodies prior to any osteophyte formation ^(50,51)^. Landmark placement is similar to that illustrated in Fig 3 of Genant et al ^(52)^. The posterior superior landmarks are placed so as to represent the endplate as it would appear on a mid-sagittal slice of a CT exam, rather than up at the top of the posterior ridges on the upper endplate similar to logic described in Keynan et al. ^(37)^. When the X-ray beam path through a vertebra is not perpendicular to the mid-sagittal plane of the vertebra, the left and right sides of the vertebral endplates and the left and right aspects of the posterior vertebral body may be seen on the X-ray.^(16,20)^ It is understood that the radiographic shadows that might be interpreted as the left and right rims of the endplates might not exactly represent the endplate rims^(16)^, but it is assumed that the line bisecting these shadows is the best-available radiographic approximation of the mid-sagittal plane. Therefore, the mid-sagittal plane is assumed to be midway between the radiographic shadows of the left and right sides of endplate, and midway between the radiographic shadows of the left and right aspects of the posterior wall. Multiple examples of landmark placement are provided online^3^ to help readers appreciate the nuances of how landmarks were placed. Vertebral morphometry calculated from landmarks is dependent on details of landmark placement, and it is therefore important to appreciate landmark placement details. The examples provided include vertebrae with abnormal morphology (as defined by the metrics presented in this paper) and help to appreciate the variety of conditions that may be identified through utilization of standardized morphology metrics.

The neural networks and coded logic produced landmark coordinates for the vertebral bodies from L1 to S1 in lumbar spine radiographs and C2 to C7 in cervical spine radiographs. An anomaly detection algorithm was then run on those data to identify anomalies due to very poor image quality, severe image artifacts, and very unusual anatomy. Those anomalies were removed from subsequent data analysis.

The coordinates of the four vertebral body corners were used to calculate six vertebral body morphology metrics:

1. anterior / posterior vertebral body height ratio (VBHR)
2. superior / inferior endplate width ratio (EPWR)
3. forward / backward diagonal ratio (FBDR)
4. height / width ratio (HWR)
5. angle between the superior and inferior endplates (EPA)
6. angle between the posterior wall and the superior endplate (PSA)

The NHANES-II lumbar radiographs were obtained with the subject in the lateral recumbent position with body flexion ^(53)^. As this is not the typical position used to obtain a radiograph for vertebral fracture detection, global and local spinal lordosis were not calculated. The NHANES-II cervical spine radiographs were obtained with the subject standing and a 25 lb weight in each hand to pull down the shoulders, and the head and neck flexed forward. ^(53)^

Principal component analysis was used to determine if the dimensionality of the data can be reduced (Stata Ver 15). The dependency of the six vertebral morphology metrics on sex, age, race, nation of origin, height, weight, and BMI was assessed using analysis of variance (ANOVA, Stata Ver 15). In the NHANES-II study, race was recorded as white, black, or other. Nation of Origin was one of 13 categories.

Descriptive statistics were tabulated and used to identify and trim outliers in the data that may compromise the definition of “normal” vertebral morphology ^(54-57)^. Outliers <> 3 standard deviations (SD) from the mean were inspected to identify typical explanations for each type of outlier. The data were trimmed by excluding all vertebrae where any of the six metrics was <> 2 SD from the mean, and descriptive statistics were tabulated for use in quantifying deviation from average for each metric. Correlations between the six morphology metrics were also explored. Finally, means and standard deviations were tabulated for each metric, for each vertebra from L1 to S1 and from C2 to C7. Those means and standard deviations were then used to calculate standardized metrics (standard deviations from mean) for the entire data sets. All statistical analysis was completed using Stata ver 15 (College Station, TX, USA).

The NHANES-II study also includes a response to the question: “Have you ever had pain in your back on most days for at least two weeks”. That data was analyzed for associations with the six morphology metrics.

To assess reproducibility of the metrics, the six morphology metrics were also calculated for flexion and extension radiographs for 415 asymptomatic volunteers. This is an expanded version of previously published data.^(58)^ The morphology metrics were calculated from landmark coordinates obtained using the same neural networks and coded logic used with the NHANES-II X-rays. The morphology of each vertebra should not have changed between flexion and extension, unless there was an unhealed vertebral fracture, which is unlikely in an asymptomatic person. However, the position of each vertebrae relative to the central beam path did change substantially between flexion and extension in most cases. The morphology measured from the flexion X-ray was compared to the morphology calculated from the extension X-ray using Bland-Altman limits of agreement. Any variability in the metrics is assumed to be error due to variability in radiographic projection.

To further document expected variability in the metrics specifically due to variations in radiographic projections that can occur in clinical practice, digitally reconstructed radiographs were used. Thin-slice (1 mm or less) anonymized computed tomography exams of the lumbar spine of 26 individuals or cadavers (from past research studies) were interpolated to 0.2 mm isotropic resolution. The three-dimensional coordinates of the four anatomic landmarks described in Figure 1 were digitized, in 3D, for the mid-sagittal plane of each vertebra from L1 to S1, using intersecting axial, coronal, and sagittal slices. Landmarks were thus placed in the mid-sagittal plane at clear and well-defined locations. This may be as close as possible to “gold standard” landmark placement. The image processing and landmark digitization was completed using Slicer 3D.^(59)^ Custom python code was developed to create 2D, digitally reconstructed radiographs (DRR) from the 3D data using Plastimatch DRR ^(60)^. The projection matrices used by Plastimatch DRR to create the X-rays were also used to calculate, from the known 3D coordinates, the precise coordinates of each landmark on the 2D simulated X-ray. A 40 inch source-to-image distance was used for all X-rays. The base x-ray was centered on the centroid of the L3 vertebra (this is the baseline isocenter). Forty additional X-rays were generated with the following variations:

- random beam tilts of between ± 10 deg in each of the sagittal and axial planes
- random displacements from the baseline isocenter ± 2 endplate widths (EPW) in the AP and LR directions, plus ± 3 EPW shifts in the cranial-caudal direction
- random image rotations between ± 24 deg

These variations resulted in a wide range of radiographic projections. The landmarks in every image were precisely known so there were zero errors in landmark coordinates relative to the 3D coordinates. This allows for documenting the effect of variability in vertebral morphology metrics that are due to radiographic projection, without uncertainty in landmark placement. These precisely calculated landmarks were analyzed to obtain the six vertebral body morphology metrics. There were thus 41 morphology measurements for every vertebra from L1 to S1 for each of 26 independent CT exams. The variability in each metric was assessed using the standardized version of the metric, since all of these metrics are thereby expressed as standard deviations from the average and that allows for combining data for all vertebrae and for consistent interpretation of the data. Although the coordinates of landmarks in every simulated X-ray were precisely calculated, the quality of the simulated radiographs was below clinical standards and not appropriate for use toward validating the accuracy of neural networks in obtaining landmarks.

## Results and Discussion

Due to the large quantity of data to be presented, the results and discussion are combined.

### Summary of Data Analyzed

Computer generated lumbar landmarks were obtained for 42,980 vertebrae from 7,364 of 7,423 total lumbar radiographs and 54,093 vertebrae from 9,662 out 9,669 cervical radiographs for subjects 25 to 74 years old. 50 lumbar X-rays were not analyzed due to an error in file transfer and seven cervical and nine lumbar X-rays were non-analyzable. Landmark generation for all lumbar and cervical radiographs took 22 hrs on a dual CPU, 4 GPU (Nvidia T4) production server. The age range was 25 to 74 years and the demographic and body size data are summarized in Table 1.

**Table 1:** Sample size, average age and [SD], and average BMI (kg/m2) and [SD]. Age and BMI were missing for two subjects.

|  | Lumbar |  |  | Cervical |  |  |
| --- | --- | --- | --- | --- | --- | --- |
| Sex | N | Age | BMI | N | Age | BMI |
| Male | 4,553 | 50.9[15.3] | 25.5[4.0] | 4,610 | 50.8[15.3] | 25.5[4.0] |
| Female | 2,809 | 63.3[6.4] | 26.4[5.5] | 5,050 | 51.4[15.2] | 25.8[5.8] |

Note that females are under-represented in the lumbar data. By design of the NHANES-II study, no lumbar X-rays were taken for pregnant women or women under 50.^(61)^ An additional limitation relates to the data on race and nation of origin recorded in the NHANES-II data. Based on the cervical data, 86.9% of NHANES-II subjects were “white”, 11.2% “black” and the rest “other”. A more uniform representation of races would likely be needed to fully understand the importance of race. The same is true for the “nation of origin” data in NHANES-II. NOTE: the language in quotes is as used in the NHANES-II documentation: 74.1% of subjects are classified as “OTHER EUROPE SUCH AS GERMAN, FRENCH, ENGLISH, IRISH”, 10.8% classified as “BLACK, NEGRO OR AFRO-AMERICAN”, and the rest distributed in 1 of 11 other classifications. Djoumessi et al. have reported differences in the VBHR between different nations of origin ^(56)^. It would be valuable to repeat this analysis on X-rays that better represent races and nations of origin not well represented in the NHANES-II data. The ages of the subjects (using cervical data) were also biased toward older ages (Figure 5). That may explain the relatively high rate of vertebrae with at least one abnormal morphology metric.

**Figure 5:**
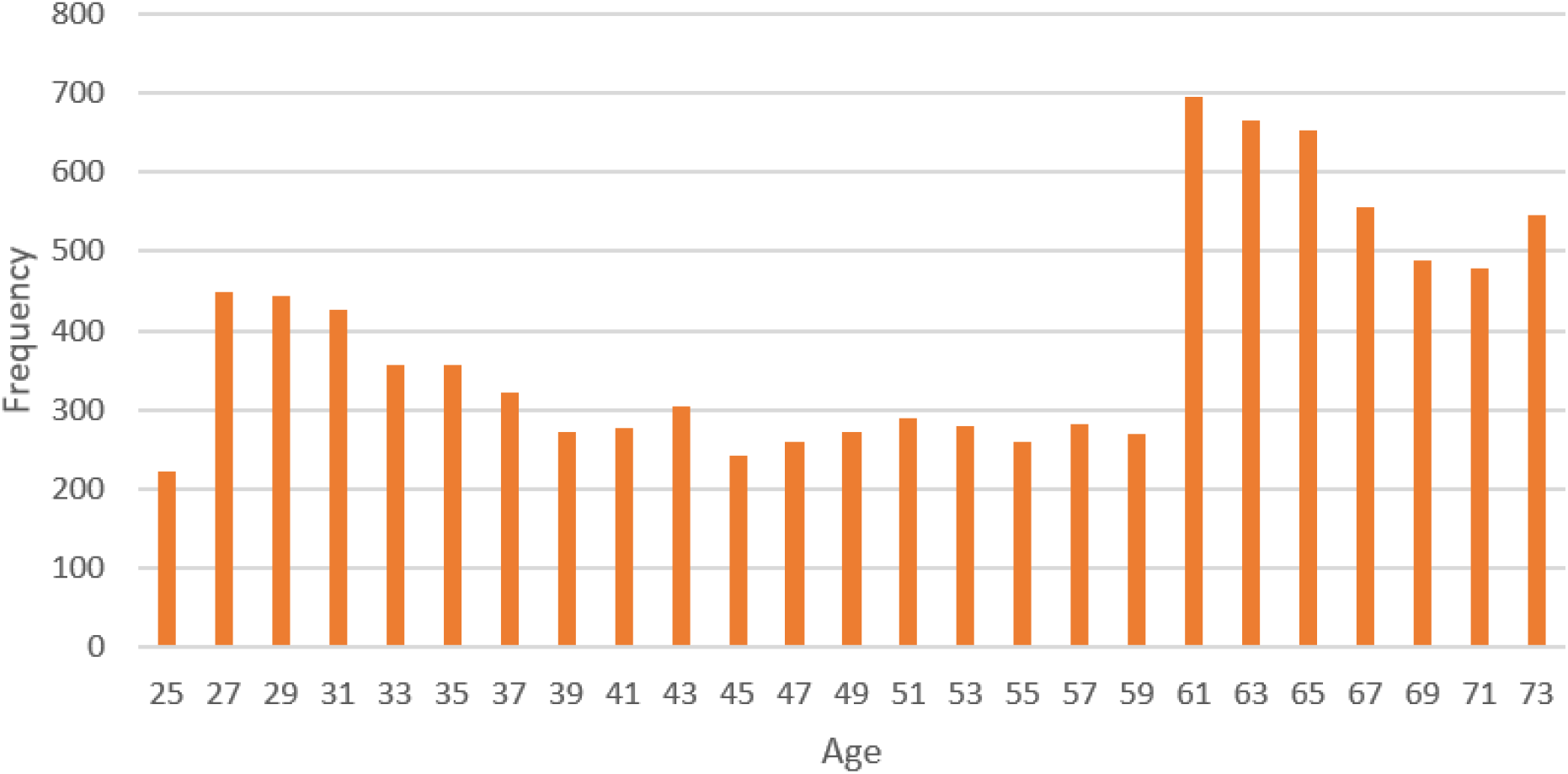
Distribution of ages in the NHANES-II study.

Principal component analysis (PCA) of the lumbar data documented that 5 primary components can explain 99.8% of the variance in the morphology data. PCA of the cervical data documents that 6 components explain 99.7% of the variance. Since PCA does not appreciably reduce the dimensionality of the data, it was not investigated further.

### Effect of covariates

Analyses of variance (after excluding outliers) revealed that age, sex, race, and nation of origin were statistically significant for most of the variables (Tables 2 and 3), and the R^2^ was high. However, the F-statistic was generally small for all variables except vertebral level. Excluding all variables except vertebra changed the R^2^ very little (e.g. for the lumbar data, VBHR R^2^ went from 0.848 to 0.842 and HWR R^2^ went from 0.839 to 0.800). This documents that age, sex, race and body size are significant, but contribute relatively little to the variability in the morphology metrics. Therefore, except for vertebral level, all data were pooled for subsequent analysis. Previous studies have shown a dependence of vertebral morphology on age, gender, race and other variables, but the relative magnitude of the influence was not described.^(62,63)^

**Table 2:** Results of multivariate analysis of variance to determine the importance of multiple independent variables for explaining variability in the lumbar vertebral morphology metrics. The “F” columns provide the “F” statistic. The “P>F” columns provide the statistical significance. The F values document how much of the variability in the dependent variable is explained by each independent variable.

| Variable | VBHR R<sup>2</sup> = 0.85 |  | EPWR R<sup>2</sup> = 0.92 |  | FBDR R<sup>2</sup> = 0.93 |  | HWR R<sup>2</sup> = 0.84 |  | EPA R<sup>2</sup> = 0.82 |  | PSA R<sup>2</sup> = 0.75 |  |
| --- | --- | --- | --- | --- | --- | --- | --- | --- | --- | --- | --- | --- |
|  | F | P > F | F | P > F | F | P > F | F | P > F | F | P > F | F | P > F |
| *Vertebra* | 39009 | **0.000** | 85907 | **0.000** | 94108 | **0.000** | 34955 | **0.000** | 32422 | **0.000** | 20937 | **0.000** |
| Age | 54 | **0.000** | 133 | **0.000** | 72 | **0.000** | 1414 | **0.000** | 8 | **0.004** | 10 | **0.002** |
| Sex | 350 | **0.000** | 82 | **0.000** | 4 | **0.054** | 3969 | **0.000** | 535 | **0.000** | 146 | **0.000** |
| Race | 7 | **0.001** | 0 | 0.716 | 2 | 0.218 | 6 | **0.002** | 8 | **0.000** | 4 | **0.021** |
| Nation of Origin | 5 | **0.000** | 1 | 0.398 | 2 | **0.034** | 13 | **0.000** | 6 | **0.000** | 4 | **0.000** |
| Weight | 0 | 0.995 | 3 | 0.089 | 0 | 0.857 | 10 | **0.002** | 0 | 0.760 | 0 | 0.506 |
| Height | 0 | 0.829 | 7 | **0.010** | 2 | 0.211 | 2 | 0.194 | 1 | 0.458 | 0 | 0.526 |
| BMI | 0 | 0.645 | 3 | 0.076 | 0 | 0.868 | 0 | 0.482 | 0 | 0.911 | 0 | 0.496 |

**Table 3:** Results of multivariate analysis of variance to determine the importance of multiple independent variables for explaining variability in the cervical vertebral morphology metrics.

| Variable | VBHR R<sup>2</sup>=0.72 |  | EPWR R<sup>2</sup>=0.77 |  | FBDR R<sup>2</sup>=0.68 |  | HWR R<sup>2</sup>=0.88 |  | EPA R<sup>2</sup>=0.72 |  | PSA R<sup>2</sup>=0.55 |  |
| --- | --- | --- | --- | --- | --- | --- | --- | --- | --- | --- | --- | --- |
|  | F | P > F | F | P > F | F | P > F | F | P > F | F | P > F | F | P > F |
| Vertebra | 22659 | **0.000** | 30247 | **0.000** | 19163 | **0.000** | 66488 | **0.000** | 22852 | **0.000** | 10977 | **0.000** |
| Age | 136 | **0.000** | 161 | **0.000** | 421 | **0.000** | 2683 | **0.000** | 103 | **0.000** | 415 | **0.000** |
| Sex | 0 | 0.553 | 14 | **0.000** | 381 | **0.000** | 829 | **0.000** | 3 | 0.065 | 236 | **0.000** |
| Race | 7 | **0.001** | 0 | 0.928 | 0 | 0.895 | 27 | **0.000** | 7 | **0.001** | 1 | 0.472 |
| Nation of Origin | 2 | **0.017** | 1 | 0.223 | 7 | **0.000** | 21 | **0.000** | 2 | **0.039** | 6 | **0.000** |
| Weight | 18 | **0.000** | 5 | **0.029** | 0 | 0.958 | 4 | **0.044** | 24 | **0.000** | 1 | 0.303 |
| Height | 27 | **0.000** | 6 | **0.017** | 25 | **0.000** | 51 | **0.000** | 34 | **0.000** | 32 | **0.000** |
| BMI | 14 | **0.000** | 5 | **0.033** | 0 | 0.752 | 0 | 0.528 | 18 | **0.000** | 0 | 0.606 |

### Descriptive Statistics for Normative Reference

Descriptive statistics for the morphologic parameters are provided in Tables 4 and 5. Mean values changed very little for the morphology metrics after removing the outliers (<> 2 SD from mean), suggesting that normal morphology is dominant in the data. Standard deviations were reduced after removing outliers. These observations are the same as reported in a similar study ^(56)^. Data with a Gaussian distribution have a skewness of 0 and kurtosis of 3. Excluding outliers substantially reduced the skewness from an average of 0.27 to 0.05, and substantially narrowed the tails of the distribution from an average kurtosis of 4.1 to 2.5. This documents that the distribution of data was more Gaussian after removing outliers. There was remarkably small variation in vertebral morphology after accounting for vertebra (L1, L2, etc.). The PSA, FBDR, and EPWR generally had the lowest coefficients of variation (Tables 6 and 7).

**Table 4:** Means and [standard deviations] for 35,275 lumbar vertebrae. All vertebrae where any metric was <> 2 Std Dev from the average for all vertebrae in the NHANES-II study were excluded.

| Vert | VBHR | EPWR | FBDR | HWR | EPA | PSA |
| --- | --- | --- | --- | --- | --- | --- |
| L1 | 0.944[0.043] | 0.994[0.017] | 0.994[0.017] | 0.823[0.059] | -2.678[2.035] | 88.499[1.250] |
| L2 | 0.979[0.040] | 0.999[0.018] | 0.973[0.019] | 0.827[0.059] | -1.021[1.907] | 87.919[1.251] |
| L3 | 0.997[0.039] | 1.009[0.017] | 0.960[0.016] | 0.821[0.058] | -0.126[1.825] | 87.222[1.266] |
| L4 | 1.032[0.046] | 1.003[0.019] | 0.938[0.017] | 0.799[0.058] | 1.406[2.049] | 86.753[1.383] |
| L5 | 1.154[0.066] | 1.023[0.025] | 0.930[0.023] | 0.788[0.058] | 6.517[2.655] | 87.671[1.970] |
| S1 | 1.296[0.071] | 1.611[0.154] | 1.268[0.067] | 1.157[0.086] | 11.937[3.456] | 96.564[3.539] |

**Table 5:**
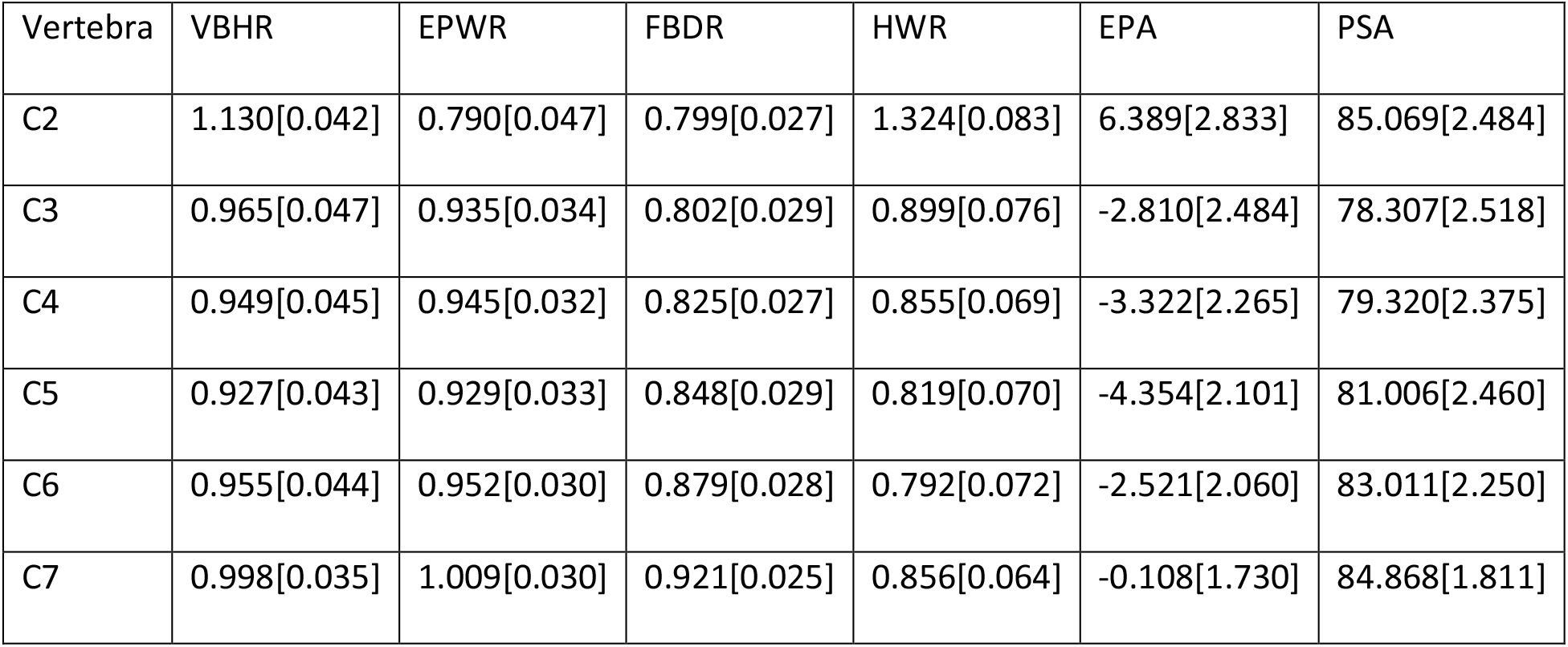
Means and [standard deviations] for 44938 cervical vertebrae. All vertebrae where any metric was <> 2 Std Dev from the average for all vertebrae in the NHANES-II study were excluded.

**Table 6:**
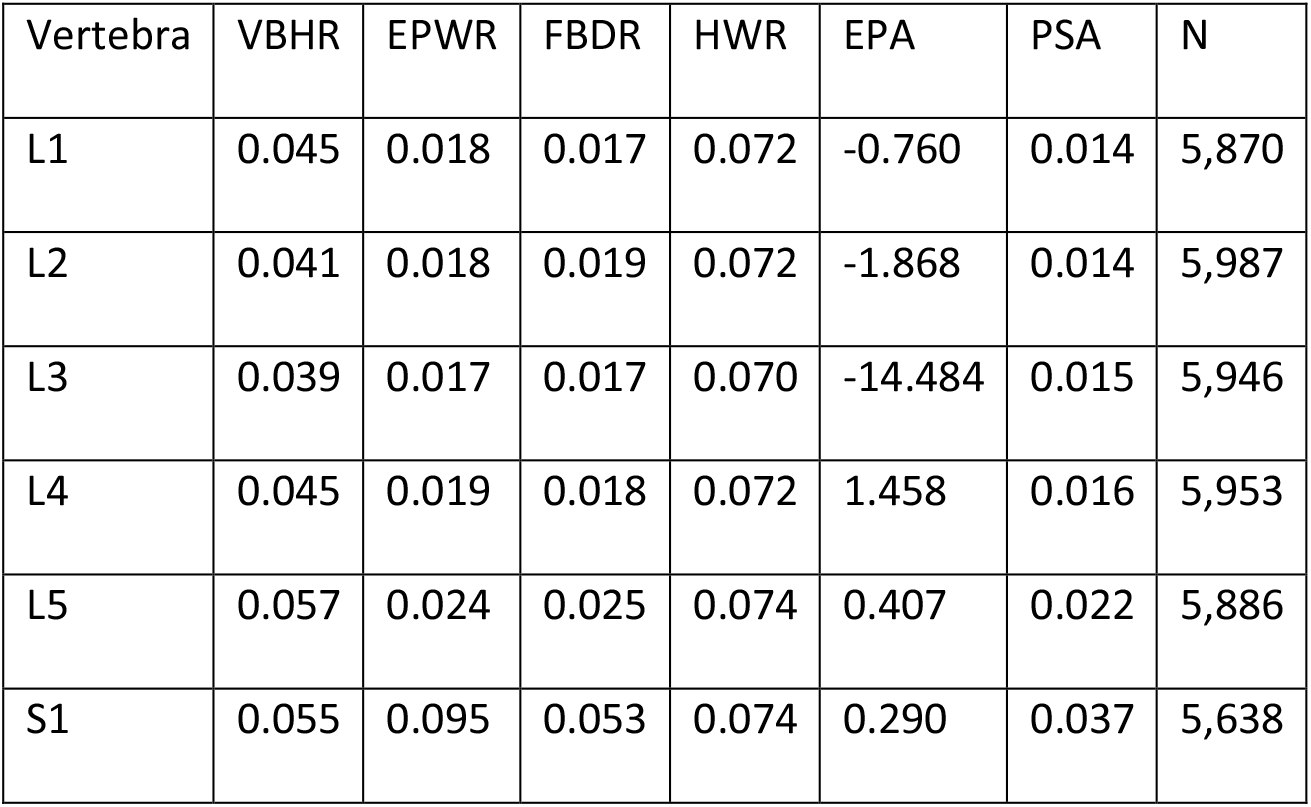
Coefficients of variation (CV = SD / mean) for 35,275 lumbar vertebrae. Note that the large values for L3 EPA is due to the mean value being very close to zero (ref Table 4) and the CV is difficult to interpret when the denominator is near zero.

**Table 7:**
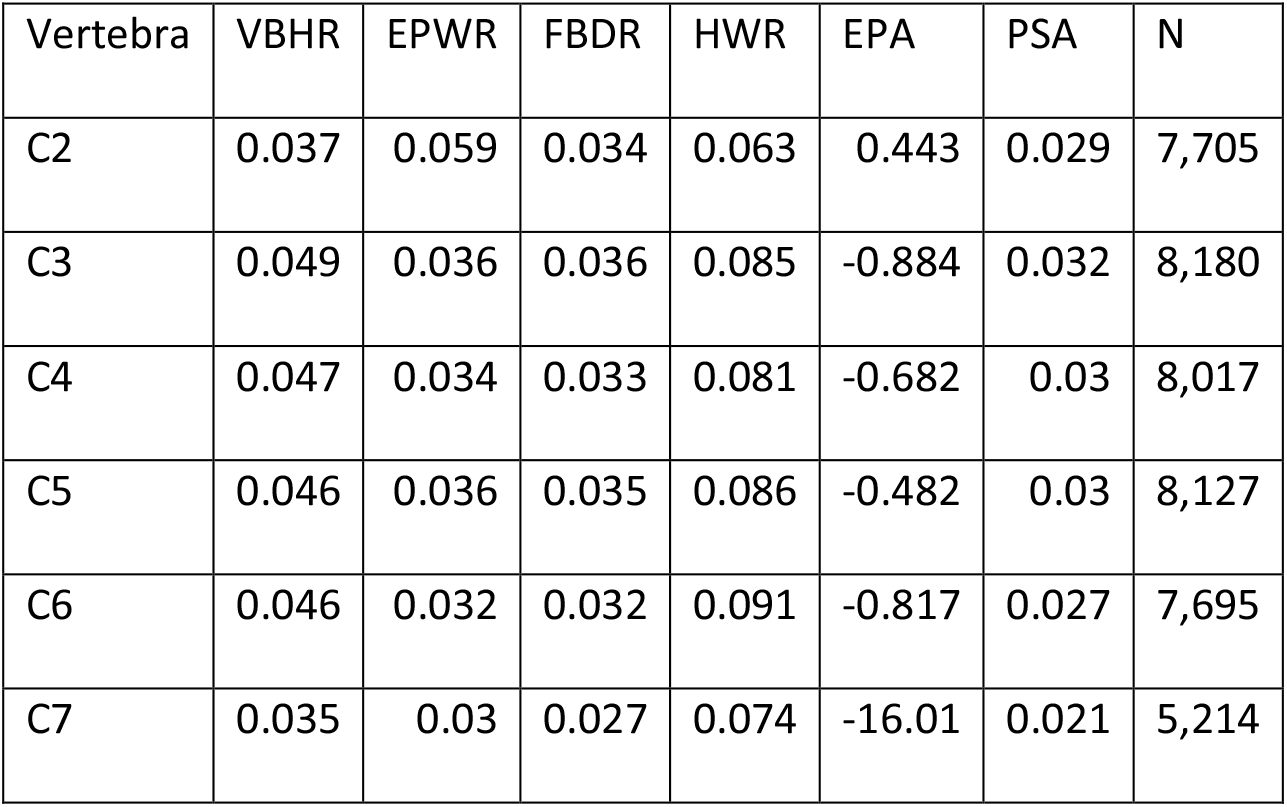
Coefficients of variation (SD / mean) for 44,938 cervical vertebrae.

Figure 6 provides comparison of VBHR data in Table 4 to some previously reported morphology metrics.^(54-57,64-66)^

**Figure 6:**
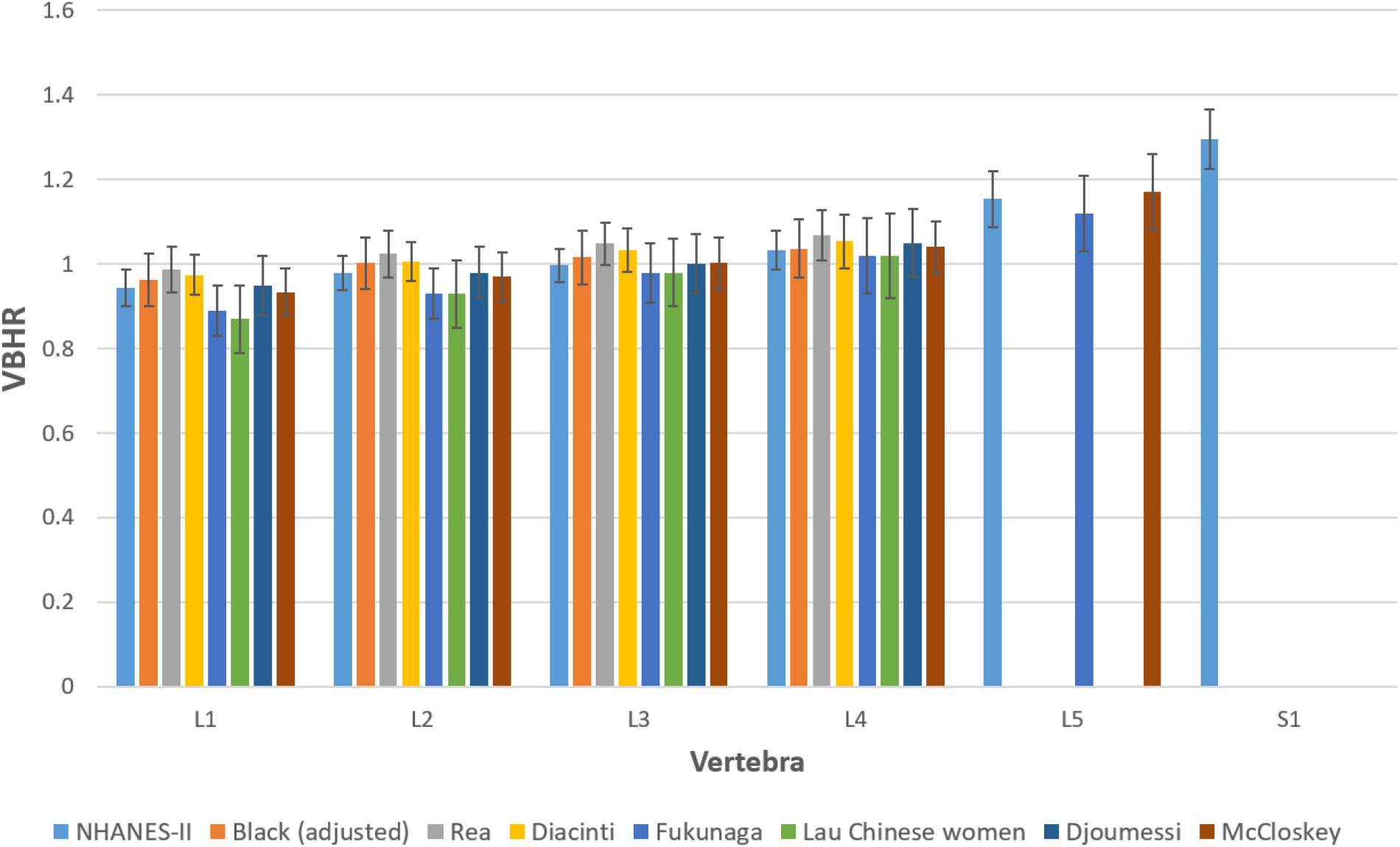
Comparison of previously published ^(54,55,57,64-66)^ VBHR data to data obtained from the NHANES-II study.

The six morphologic variables were significantly correlated with each other (Tables 8 and 9). EPA and VBHR were almost perfectly correlated, suggesting that only one of these two variables needs to be analyzed. Although VBHR and EPA were almost perfectly correlated, the other metrics were not. That observation supports that the metrics each independently describe some unique aspect of vertebral morphology. It cannot be determined from the NHANES data alone which of the six metrics will prove most valuable in research and clinical practice. It may be worth exploring the utility of each metric, and combinations of metrics, in future studies where the goal is to diagnose fractures or deformities, or to predict response to treatment.

**Table 8:** Pearson’s correlation coefficients between morphologic variables for 35,275 lumbar vertebrae, after excluding vertebrae where any of the morphologic variables was ±2 SD from the average for 42,980 vertebrae.

|  | VBHR | EPWR | FBDR | HWR | EPA | PSA |
| --- | --- | --- | --- | --- | --- | --- |
| VBHR | 1 |  |  |  |  |  |
| EPWR | 0.7741 | 1 |  |  |  |  |
| FBDR | 0.6694 | 0.9373 | 1 |  |  |  |
| HWR | 0.6661 | 0.8844 | 0.8712 | 1 |  |  |
| EPA | 0.9904 | 0.7338 | 0.6205 | 0.6551 | 1 |  |
| PSA | 0.7394 | 0.7911 | 0.9069 | 0.8069 | 0.7188 | 1 |

**Table 9:** Pearson’s correlation coefficients between morphologic variables for 44,938 cervical vertebrae, after excluding vertebrae where any of the morphologic variables was ±2 SD from the average for 54,093 vertebrae.

|  | VBHR | EPWR | FBDR | HWR | EPA | PSA |
| --- | --- | --- | --- | --- | --- | --- |
| VBHR | 1 |  |  |  |  |  |
| EPWR | -0.6536 | 1 |  |  |  |  |
| FBDR | -0.2641 | 0.5298 | 1 |  |  |  |
| HWR | 0.7861 | -0.7548 | -0.4464 | 1 |  |  |
| EPA | 0.9848 | -0.5813 | -0.1828 | 0.7646 | 1 |  |
| PSA | 0.5704 | -0.352 | 0.5461 | 0.3203 | 0.6047 | 1 |

Multiple prior publications report efforts to establish reference data that can be used to help diagnose vertebral body fractures. ^(6-10,12-15,17-43)^ Although multiple metrics need to be considered, such as intervertebral angles, listhesis and canal occlusion ^(37)^, compression of the vertebral body height is commonly used to identify fractures. When severe loss of vertebral body height has occurred, clinicians can unanimously agree that a fracture has occurred, but when the fracture is subtle, reference data may be important. Mid-vertebral landmarks that could be used to measure mid-vertebral height were not routinely available in the images used to train the neural networks, so mid-vertebral body heights could not be obtained automatically. Change in vertebral shape is considered one of the best indicators of a fracture ^(6)^. The NHANES-II images provide only one radiograph per individual, so change in shape cannot be explored. Nevertheless, the low variability of vertebral shape within a large population supports the potential for using a single X-ray to detect abnormal morphology with reference to the NHANES-II data. It is also likely, that with advancements in the application of artificial intelligence to producing spine metrics, the data in Tables 4 and 5 may provide valuable quality control data to determine if the AI predictions are aberrant or if caution is needed in interpreting the results.

Application of the vertebral morphology metrics, such as in Tables 4 and 5, to the challenge of documenting vertebral fractures will require defining a threshold level of these metrics that can be used to document the presence/absence of a fracture. Eastell et al. and McCloskey et al. used a threshold of 3 SD for the VBHR ^(64,67)^. Black et al. used a threshold of 20% difference from normal VBHR ^(25)^. Genant et al. used the criteria of a 15% difference compared to the mean VBHR of a normal population ^(52)^. The average VBHR in Table 4 for L1 to L4 is about 1 and one SD is about 0.042, so a 15% difference as used by Genant et al. would be equivalent to approximately a VBHR Z-score of 3.5. A gold-standard test for a true vertebral fracture does not exist, so validating a threshold remains a challenge. An alternative, treatment specific approach, would be to validate a threshold that predicts a positive response to treatment. That approach could potentially be retrospectively tested at low cost using automated measurements of morphology applied to imaging and data from prior studies that collected appropriate imaging.

### Differences between levels

It is reasonable to hypothesize that differences in morphology between adjacent levels may help to identify abnormal morphology. For example, if one level has an anterior wedge fracture, then a significant difference in the VBHR would be expected when compared to the immediately adjacent levels. A problem occurs when adjacent vertebrae are both fractured or otherwise abnormal. Comparison to adjacent levels has been a criteria used or proposed by many investigators, so data that could be used to implement adjacent vertebrae comparisons are provided in tables 10 and 11. For example, using data in Table 10, if the VBHR is to be compared between L4 and L5 vertebrae, and normal variability is defined as anything within the 95% confidence interval for data from the NHANES-II study, than the VBHR when comparing L4 to L5 should not vary by more than 0.124 ± 1.96*0.059, or 0.007 to 0.24.

**Table 10:** The means and [standard deviations] for the absolute differences between adjacent lumbar vertebrae for each of the vertebral morphology metrics. These data include only those vertebrae with no abnormalities in any morphology metric using the data in Table 4. The sample size is smaller, since if either vertebra had any abnormality, that comparison between levels was excluded.

| Compare | N | VBHR | EPWR | FBDR | HWR | EPA | PSA |
| --- | --- | --- | --- | --- | --- | --- | --- |
| L1 to L2 | 3540 | 0.041[0.030] | 0.017[0.013] | 0.024[0.016] | 0.023[0.018] | 2.009[1.442] | 1.127[0.864] |
| L2 to L3 | 3635 | 0.034[0.026] | 0.018[0.014] | 0.018[0.013] | 0.023[0.019] | 1.611[1.224] | 1.136[0.847] |
| L3 to L4 | 3628 | 0.043[0.031] | 0.018[0.013] | 0.025[0.015] | 0.028[0.021] | 1.955[1.410] | 1.106[0.838] |
| L4 to L5 | 3554 | 0.124[0.059] | 0.026[0.019] | 0.020[0.015] | 0.027[0.022] | 5.191[2.443] | 1.583[1.178] |
| L5 to S1 | 3137 | 0.145[0.080] | 0.589[0.134] | 0.342[0.061] | 0.372[0.074] | 5.733[3.542] | 9.113[3.504] |

**Table 11:** The means and [standard deviations] for the absolute differences between adjacent cervical vertebrae for each of the vertebral morphology metrics. These data include only those vertebrae with no abnormalities in any morphology metric using the data in Table 5.

| Compare | N | VBHR | EPWR | FBDR | HWR | EPA | PSA |
| --- | --- | --- | --- | --- | --- | --- | --- |
| C2 to C3 | 4636 | 0.167[0.053] | 0.145[0.053] | 0.025[0.018] | 0.426[0.070] | 9.297[3.241] | 6.736[2.868] |
| C3 to C4 | 5039 | 0.041[0.031] | 0.032[0.024] | 0.027[0.019] | 0.052[0.038] | 2.063[1.553] | 1.983[1.475] |
| C4 to C5 | 5023 | 0.041[0.031] | 0.033[0.025] | 0.028[0.020] | 0.047[0.034] | 2.046[1.509] | 2.207[1.607] |
| C5 to C6 | 4850 | 0.045[0.033] | 0.036[0.026] | 0.034[0.022] | 0.044[0.034] | 2.429[1.732] | 2.466[1.719] |
| C6C7 | 3231 | 0.051[0.035] | 0.060[0.036] | 0.045[0.025] | 0.064[0.040] | 2.713[1.768] | 2.363[1.619] |

### Abnormalities in the NHANES data

Mean and standard deviation data from Table 4 and Table 5 were used to calculate the vertebra specific Z-score for each metric according to the following equation

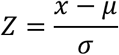

where *x* is the metric for a vertebra, *μ* is the vertebra specific mean for the corresponding metric, and *σ* is the vertebra specific standard deviation for the corresponding metric. Based on these Z-scores, 28,813 of 42,980 lumbar vertebrae (67%) were within ± 2 SD of average (Table 4) for all six of the metrics. These number are less than obtained after trimming since the standard deviations in Table 4 are smaller than originally used to identify outliers, and are therefore more sensitive to abnormalities. Similarly, 36,869 of 54,093 cervical vertebrae (68%) were within ± 2 SD of average (Table 5) for all six of the metrics.

Box and whisker plots illustrate the magnitude of outliers that exist in the NHANES data (Figures 7 and 8). Note that the medians are generally centered within the interquartile range and the whiskers are symmetric. Positive VBHR outliers represent posterior wedge-shaped vertebral bodies, while negative VBHR outliers represent anterior wedge-shaped vertebrae. The wedging may either be from fractures, remodeling, or congenital conditions and identifying a specific cause would require more than a single radiograph. Positive EPWR outliers represent vertebrae that are unusually wide superiorly versus inferiorly, and negative outliers are the opposite. Positive and negative FBDR outliers generally appear to be best explained by remodeling, perhaps after a fracture that caused splaying of the fracture fragments. Positive HWR outliers appear as unusually tall vertebrae, while negative HWR outliers generally appear to be best explained by a crush fracture. Positive EPA outliers appear as anteriorly wedge-shaped vertebrae while negative outliers appear as posteriorly wedge-shaped vertebrae. Positive PSA outliers generally appear as posterior wedge-shaped vertebrae and negative outliers generally appear as anteriorly wedge-shaped vertebrae. It is not possible to definitively identify the cause of each abnormality from a single radiograph. Examples of each of high positive and low negative outliers, for each of the six metrics, including landmarks, can be viewed online. (https://www.dropbox.com/sh/qzrocrh86goxarx/AAADRXs5HoEjVdRcGIwA4beIa?dl=0).

**Figure 7:**
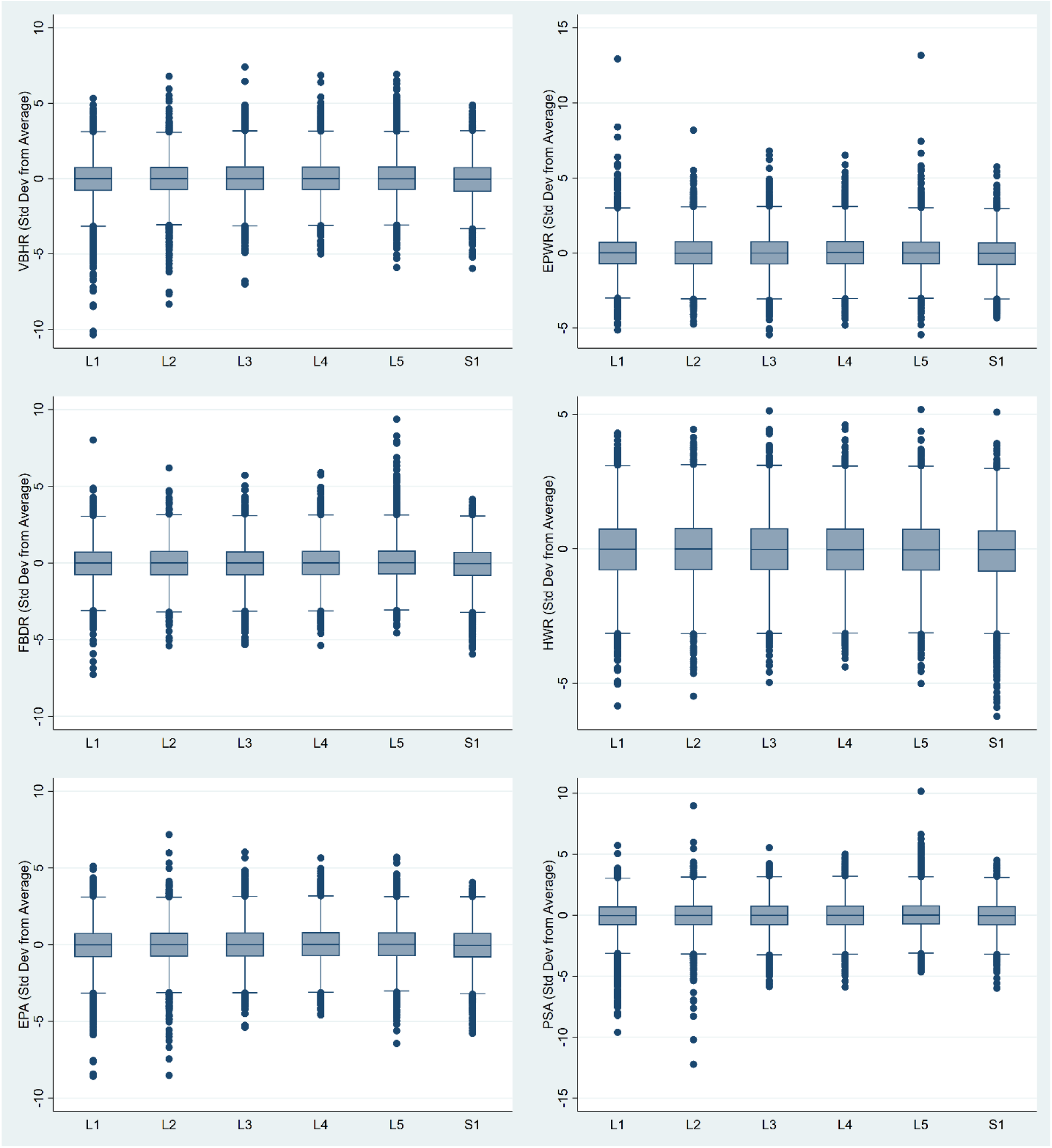
Box and whisker plots for the six morphology metrics calculated for 42,980 lumbar vertebrae. These plots document the types and magnitude of outliers in the data.

**Figure 8:**
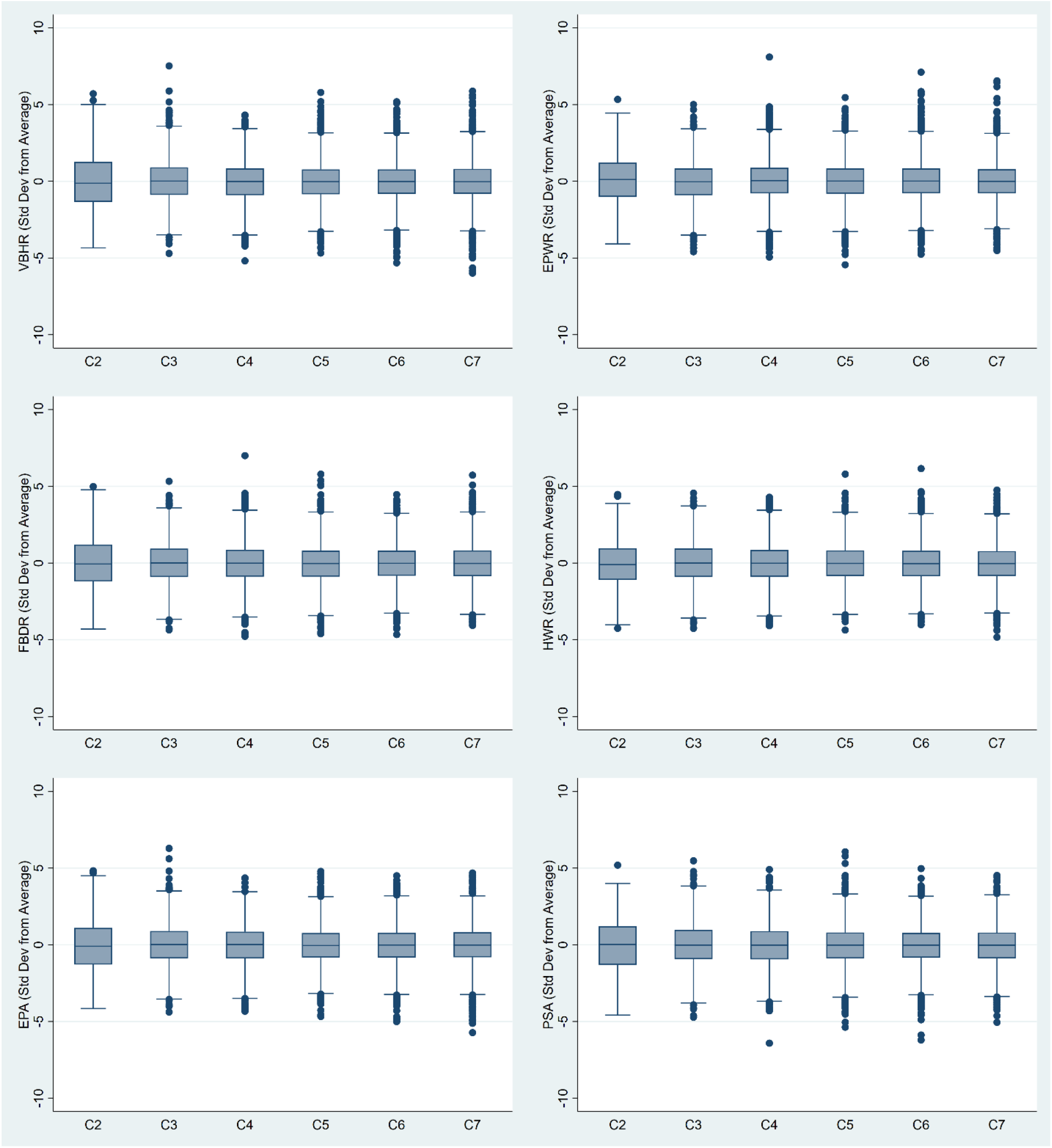
Box and whisker plots for the six morphology metrics calculated for 54,093 cervical vertebrae. These plots document the types and magnitude of outliers in the data.

### Error analysis for the morphology metrics

One advantage of the six vertebral morphology metrics is the lack of dependence on radiographic magnification. All metrics are ratios of single vertebra dimensions or angles between anatomic reference lines and should not vary appreciably with radiographic magnification (assuming sufficient spatial resolution). This is important since radiographic magnification can vary substantially between lumbar X-rays ^(78-80)^ and this can result in errors in measurements of vertebral dimensions ^(26)^.

Some error in the data from Tables 4 and 5 may occur as a result of the challenges of vertebral labeling. A systematic approach to definitively labeling vertebrae requires whole spine imaging ^(68)^. An X-ray or CT of the entire spine from occiput to sacrum was not available (X-rays typically included T10 to S1), so it cannot be certain that vertebrae were optimally labeled if there were <> 5 lumbar vertebrae transitional vertebrae, or other abnormalities ^(69,70)^. Spines with <> 5 lumbar vertebrae are uncommon^(71)^. Transitional vertebrae may be more common^(72)^, but it is unclear whether this would consistently compromise labeling from a lateral spine x-ray. The assumption is that the vertebrae were correctly labeled in the majority of cases and the possibility of mislabeled vertebrae in a minority of cases would not significantly affect the results. The prevalence of transitional vertebrae reported in the peer-reviewed literature varies between 4 and 36% ^(73-77)^. The prevalence in the NHANES-II radiographs is unknown.

The six morphology metrics calculated from flexion X-rays were compared to the metrics calculated from paired extension X-rays for 2,132 vertebrae from L1 to S1. There should be no difference in vertebral morphology between flexion and extension (assuming no mobile vertebral body fractures). Table 12 provides the results of the Bland-Altman limits of agreement analysis. This variability is assumed due to the differences in radiographic projection between images. Typically, there would not be much difference in radiographic projection between carefully obtained flexion versus extension X-rays, and this is reflected in the mean differences in Table 12, but substantial differences did occur in some subjects, and this is reflected in the low and high limits of agreement in Table 12. This experiment documents that some morphology metrics could vary by as much as 1.65 Std Dev due to radiographic projection differences. Thus, it is important to strive for radiographs where the vertebrae of interest are positioned near the center of the radiograph. The experiment with the simulated X-rays, discussed below, compliments and helps to explain results in Table 12.

**Table 12:** Results of a Bland-Altman limits of agreement analysis for morphology (expressed as SD from the average) measured from 2,132 lumbar flexion X-rays compared to paired extension X-rays. The mean differences between the two measurements, as well as the low and high limits of agreement are provided. These data are specific to the methods used to obtain anatomic landmarks from the radiographs.

| Metric | Mean<br>Difference | Low | High |
| --- | --- | --- | --- |
| VBHR | 0.15 | -0.93 | 1.24 |
| EPWR | 0.028 | -1.59 | 1.65 |
| FBDR | -0.09 | -1.58 | 1.4 |
| HWR | -0.016 | -0.92 | 0.89 |
| EPA | 0.16 | -0.99 | 1.3 |
| PSA | 0.02 | -1.57 | 1.61 |

### Variability due to radiographic projection

The experiment using simulated X-rays was completed to help understand how variability in radiographic projection can affect the vertebral morphology metrics. This could be important, for example, with sub-optimally positioned patients, or in patients with scoliosis or other deformities, all of which can result in the x-ray beam passing from source to image obliquely to the vertebral endplates or the posterior wall of the vertebrae. There should ideally be no variability in vertebral morphology in the experiment using the simulated x-rays, since all x-rays (for a subject) were created from the same CT exam. All landmarks were precisely calculated from one set of 3D landmark coordinates, based on the geometry of the radiographic projection. Any variability is due to the effect of variability in radiographic projection on calculation of morphology. Variability was assessed using the standardized version of the metrics so that all metrics have the same units (standard deviations from average) and can thus be more easily compared. Conventional measures of variability, such as the coefficient of variation (CV = std dev / mean) or the coefficient of quartile variation (CQV = 100* (Q_3_ – Q_1_)/ (Q_3_ + Q_1_)) were not used as the mean values and (Q_3_ + Q_1_) were close to zero for some of the standardized metrics and CV or QCV are uninterpretable when the denominator approaches zero. Instead, the median and the range (max – min) for the standardized metrics was calculated for the 41 simulated X-rays that were generated from each CT exam. The median and 95^th^ percentile for these ranges, across the 26 CT exams, are provided in Table 13. In Table 13, the observed ranges of standardized metrics due to variability in radiographic projection was largest for FBDR and PSA, although the experiment demonstrates that all of the morphology metrics can vary by over 1 std dev, solely due to variability in radiographic projection. This is not as much a weakness in the metrics as it is evidence to use caution when interpreting these metrics in the presence of substantial out-of-plane imaging. It is of course possible that strategies can be developed for mitigating the effect of variability in radiographic projection. One option would be a neural network that determines the type and magnitude of out-of-plane and uses that data to correct the morphology metrics. It is assumed that the error sources discussed above were random sources of error and would not appreciably affect the means and standard deviations in Table 4 and Table 5.

**Table 13:** Variability in standardized metrics (Std Dev from Average Normal) due to variability in radiographic projection. Each of the morphology metrics was calculated for each of the 41 radiographic projections generated from each CT exam. The range in each metric (5^th^ to 95^th^ percentile) was then calculated over all 41 random variations in radiographic projection. This table has the median and 95^th^ percentile variability over the 26 CT exams. For example, due to a wide range in radiographic projections, EPA will typically vary by 0.37 Std Devs but could vary by as much as 1.1 Std Devs from the average EPA in a normal vertebra.

| Metric | Median | 95<sup>th</sup><br>Percentile |
| --- | --- | --- |
| EPA | 0.3658 | 1.0796 |
| EPWR | 0.6643 | 2.2362 |
| FBDR | 2.4421 | 4.1485 |
| HWR | 0.6378 | 1.1342 |
| PSA | 2.1198 | 3.7636 |
| VBHR | 0.3628 | 1.2465 |

### Additional observations

Based on logistic regression, the subject response to the question: “Have you ever had pain in your back on most days for at least two weeks” was significantly (P<0.05) associated with age, race, and BMI. Since there is no way to determine which vertebra (if any) might be causing symptoms, the sum of abnormalities for each subject (metrics < -2 or > 2 SD from the average based on Table 4) from L1 to S1 was calculated. This sum of abnormalities for the EPWR and the FBDR was significantly associated with the response to the “ever had back pain” question (P<0.05), although the R^2^ for these regressions was very small (>0.01), indicating a very weak association.

Of the six morphology metrics, FBDR was the most sensitive and specific metric (area under ROC curve=0.995) to use for the purpose of using landmarks to classify vertebrae as S1 vs L1-L5. A threshold level of FBDR>1.0574 misclassified only 278/42980 vertebrae as S1 vs L1-L5. Including all data for all vertebrae, the proportions of vertebrae with abnormal morphology (<> 2 SD from average based on Tables 4 and 5) are summarized in Tables 14 and 15.

**Table 14:** Percent of lumbar vertebrae in the NHANES-II study where the morphology metric was either > 2 or < -2 SD from the average, using data from Table 4 for the average and SD, and including all vertebrae.

| Vert | Freq | VBHR |  | EPWR |  | FBDR |  | HWR |  | EPA |  | PSA |  |
| --- | --- | --- | --- | --- | --- | --- | --- | --- | --- | --- | --- | --- | --- |
|  |  | < -2 | > 2 | < -2 | > 2 | < -2 | > 2 | < -2 | > 2 | < -2 | > 2 | < -2 | > 2 |
| L1 | 7,037 | 7.1 | 3.3 | 3.9 | 4.9 | 4.8 | 4.0 | 4.9 | 4.0 | 6.5 | 3.5 | 6.8 | 3.6 |
| L2 | 7,324 | 5.5 | 4.8 | 3.9 | 5.0 | 4.9 | 4.2 | 4.6 | 4.3 | 5.1 | 4.9 | 5.5 | 4.3 |
| L3 | 7,318 | 4.5 | 5.8 | 3.9 | 5.1 | 5.2 | 4.2 | 4.5 | 4.3 | 4.2 | 5.6 | 5.4 | 4.8 |
| L4 | 7,333 | 3.6 | 5.8 | 3.8 | 5.4 | 4.5 | 4.4 | 4.7 | 4.0 | 3.9 | 5.4 | 5.0 | 5.0 |
| L5 | 7,181 | 3.9 | 5.9 | 3.5 | 5.4 | 3.2 | 6.1 | 4.8 | 4.1 | 4.4 | 5.4 | 4.4 | 6.4 |
| S1 | 6,787 | 6.1 | 4.6 | 8.7 | 3.8 | 8.7 | 3.8 | 8.1 | 3.0 | 5.5 | 3.9 | 5.5 | 4.0 |
| All | 42,980 | 5.1 | 5.0 | 4.6 | 4.9 | 5.2 | 4.5 | 5.2 | 4.0 | 4.9 | 4.8 | 5.4 | 4.7 |

**Table 15:** Percent of lumbar vertebrae in the NHANES-II study where the morphology metric was either > 2 or < -2 SD from the average, using data from Table 5 for the average and SD, and including all vertebrae.

| Vert | Freq | VBHR |  | EPWR |  | FBDR |  | HWR |  | EPA |  | PSA |  |
| --- | --- | --- | --- | --- | --- | --- | --- | --- | --- | --- | --- | --- | --- |
|  |  | < -2 | > 2 | < -2 | > 2 | < -2 | > 2 | < -2 | > 2 | < -2 | > 2 | < -2 | > 2 |
| C2 | 9,647 | 4.9 | 5.3 | 3.8 | 5.1 | 4.3 | 5.3 | 4.4 | 4.1 | 4.9 | 4.8 | 5.3 | 4.3 |
| C3 | 9,648 | 3.8 | 4.5 | 4.9 | 3.7 | 3.8 | 4.2 | 4.0 | 4.2 | 3.7 | 4.1 | 3.7 | 4.5 |
| C4 | 9,654 | 4.6 | 4.3 | 4.2 | 4.5 | 3.9 | 4.5 | 4.4 | 3.9 | 4.6 | 3.9 | 4.7 | 4.2 |
| C5 | 9,613 | 4.2 | 3.8 | 4.4 | 4.5 | 4.3 | 3.8 | 4.1 | 3.8 | 4.1 | 3.7 | 4.8 | 3.4 |
| C6 | 9,148 | 4.5 | 4.0 | 3.7 | 4.9 | 4.1 | 4.2 | 4.4 | 4.0 | 4.2 | 3.9 | 4.6 | 3.3 |
| C7 | 6,383 | 5.0 | 4.6 | 3.9 | 4.9 | 4.6 | 4.1 | 4.6 | 4.3 | 4.8 | 4.3 | 5.7 | 4.4 |
| All | 54,093 | 4.5 | 4.4 | 4.1 | 4.6 | 4.1 | 4.4 | 4.3 | 4.0 | 4.4 | 4.1 | 4.8 | 4.0 |

## Conclusion

The lateral spine lumbar and cervical radiographs from the NHANES-II study were used to establish normative vertebral morphology reference data. These data can be used to help objectively identify vertebral body fractures, with the assumption that fractures will result in significant deviations from large population-based average vertebral body dimensions. Early diagnosis of fracture allows for early evaluation and prevention ultimately decreasing deformity as well as risk of future fracture. The data in Table 4 and Table 5 can be used to produce standardized metrics in units of standard deviation from average. This allows for easy identification of abnormalities resulting from vertebral fractures, atypical vertebral body morphologies, and other congenital or degenerative conditions, and removes the effect of vertebra allowing data for all vertebrae to be pooled in research studies. Metrics developed from a large database such as NHANES-II should help with differentiating between fractures and normal variants of shape. The deterministic thresholds for standardized metrics that can provide clinically meaningful diagnostic or prognostic information are yet to be determined. These data may also be useful for quality control in technology designed to automatically obtain landmarks coordinates and other metrics from medical imaging.

## Data Availability

Vertebral morphology data produced in the present study are available upon reasonable request to the authors

https://www.dropbox.com/sh/qzrocrh86goxarx/AAADRXs5HoEjVdRcGIwA4beIa?dl=0

## Footnotes

1 https://www.n.cdc.gov/nchs/nhanes/nhanes2

2 https://www.cdc.gov/nchs/nhanes/index.htm

3 https://www.dropbox.com/sh/qzrocrh86goxarx/AAADRXs5HoEjVdRcGIwA4beIa?dl=0

## References

1. Lindsay R, Silverman SL, Cooper C, Hanley DA, Barton I, Broy SB, et al. Risk of new vertebral fracture in the year following a fracture. Jama. 2001;285(3):320–3.

2. Ross PD, Davis JW, Epstein RS, Wasnich RD. Pre-existing fractures and bone mass predict vertebral fracture incidence in women. Ann Intern Med. 1991;114:919–23.

3. Lunt M, O’Neill TW, Felsenberg D, Reeve J, Kanis JA, Cooper C, et al. Characteristics of a prevalent vertebral deformity predict subsequent vertebral fracture: results from the European Prospective Osteoporosis Study (EPOS). Bone. 2003;33(4):505–13.

4. Gehlbach S, Bigelow C, Heimisdottir M, May S, Walker M, Kirkwood J. Recognition of vertebral fracture in a clinical setting. Osteoporosis international. 2000;11(7):577–82.

5. Delmas PD, van de Langerijt L, Watts NB, Eastell R, Genant H, Grauer A, et al. Underdiagnosis of vertebral fractures is a worldwide problem: the IMPACT study. Journal of bone and mineral research. 2005;20(4):557–63.

6. Ferrar L, Jiang G, Adams J, Eastell R. Identification of vertebral fractures: an update. OsteoporosInt. 2005;16(7):717–28.

7. Veleanu C, Diaconescu N. Contribution to the clinical anatomy of the vertebral column. Considerations on the stability and the instability at the height of the “vertebral units”. Anatomischer Anzeiger. 1975;137(3):287–95.

8. Saraste H, Brostrom LA, Aparisi T, Axdorph G. Radiographic measurement of the lumbar spine. A clinical and experimental study in man. Spine. 1985;10(3):236–41.

9. Frymoyer JW, Phillips RB, Newberg AH, MacPherson BV. A comparative analysis of the interpretations of lumbar spinal radiographs by chiropractors and medical doctors. Spine. 1986;11(10):1020–3.

10. Gallagher JC, Hedlund LR, Stoner S, Meeger C. Vertebral morphometry: normative data. Bone Miner. 1988;4(2):189–96.

11. Davies KM, Recker RR, Heaney RP. Normal vertebral dimensions and normal variation in serial measurements of vertebrae. J Bone Min Res. 1989;4:341–9.

12. Smith-Bindman R, Cummings SR, Steiger P, Genant HK. A comparison of morphometric definitions of vertebral fracture. JBone MinerRes. 1991;6(1):25–34.

13. Wang TM, Shih C. Morphometric variations of the lumbar vertebrae between Chinese and Indian adults. Acta Anat(Basel). 1992;144(1):23–9.

14. Nicholson PH, Haddaway MJ, Davie MW, Evans SF. Vertebral deformity, bone mineral density, back pain and height loss in unscreened women over 50 years. Osteoporosis International. Dec 1993;3(6):300–7.

15. O’Neill T, Varlow J, Felsenberg D, Johnell O, Weber K, Marchant F, et al. Variation in vertebral height ratios in population studies. J Bone Miner Res. 1994;12:1895–907.

16. Brinckmann P, Frobin W, Biggemann M, Hilweg D, Seidel S, Burton K, et al. Quantification of overload injuries to thoracolumbar vertebrae and discs in persons exposed to heavy physical exertions or vibration at the work-place: The shape of vertebrae and intervertebral discs—study of a young, healthy population and a middle-aged control group. Clinical Biomechanics. 1994;9:S3–S83.

17. Black DM, Palermo L, Nevitt MC, Genant HK, Epstein R, San VR, et al. Comparison of methods for defining prevalent vertebral deformities: the Study of Osteoporotic Fractures. JBone MinerRes. 1995;10(6):890–902.

18. Wu CY, Li J, Jergas M, Genant HK. Comparison of semiquantitative and quantitative techniques for the assessment of prevalent and incident vertebral fractures. OsteoporosInt. 1995;5(5):354–70.

19. Genant HK, Jergas M, Palermo L, Nevitt M, Valentin RS, Black D, et al. Comparison of semiquantitative visual and quantitative morphometric assessment of prevalent and incident vertebral fractures in osteoporosis The Study of Osteoporotic Fractures Research Group. JBone MinerRes. 1996;11(7):984–96.

20. Frobin W, Brinckmann P, Biggemann M, Tillotson M, Burton K. Precision measurement of disc height, vertebral height and sagittal plane displacement from lateral radiographic views of the lumbar spine. Clinical Biomechanics. 1997;12:S1–S63.

21. Genant HK. Assessment of vertebral fractures in osteoporosis research. JRheumatol. 1997;24(6):1212–4.

22. Brinckmann P, Frobin W, Biggemann M, Tillotson M, Burton K, Burke C, et al. Quantification of overload injuries to thoracolumbar vertebrae and discs in persons exposed to heavy physical exertions or vibration at the workplace Part II Occurrence and magnitude of overload injury in exposed cohorts. Clinical Biomechanics. 1998;13:S1–S36.

23. Espeland A, Korsbrekke K, Albrektsen G, Larsen JL. Observer variation in plain radiography of the lumbosacral spine. BrJ Radiol. 1998;71(844):366–75.

24. Ferrar L, Eastell R. Identification of vertebral deformities in men: comparison of morphometric radiography and morphometric X-ray absorptiometry. OsteoporosInt. 1999;10(2):167–74.

25. Black DM, Palermo L, Nevitt MC, Genant HK, Christensen L, Cummings SR. Defining incident vertebral deformity: a prospective comparison of several approaches. The Study of Osteoporotic Fractures Research Group. JBone MinerRes. 1999;14(1):90–101.

26. Weber K, Lunt M, Gowin W, Lauermann T, Armbrecht G, Wieland E, et al. Measurement imprecision in vertebral morphometry of spinal radiographs obtained in the European Prospective Osteoporosis Study: consequences for the identification of prevalent and incident deformities. BrJ Radiol. 1999;72(862):957–66.

27. Zhou SH, McCarthy ID, McGregor AH, Coombs RR, Hughes SP. Geometrical dimensions of the lower lumbar vertebrae--analysis of data from digitised CT images. Eur Spine J. 2000;9(3):242–8.

28. Szulc P, Munoz F, Sornay-Rendu E, Paris E, Souhami E, Zanchetta J, et al. Comparison of morphometric assessment of prevalent vertebral deformities in women using different reference data. Bone. 2000;27(6):841–6.

29. Ferrar L, Jiang G, Barrington NA, Eastell R. Identification of vertebral deformities in women: comparison of radiological assessment and quantitative morphometry using morphometric radiography and morphometric X-ray absorptiometry. JBone MinerRes. 2000;15(3):575–85.

30. Rea JA, Chen MB, Li J, Blake GM, Steiger P, Genant HK, et al. Morphometric X-ray absorptiometry and morphometric radiography of the spine: a comparison of prevalent vertebral deformity identification. JBone MinerRes. 2000;15(3):564–74.

31. Rea JA, Chen MB, Li J, Marsh E, Fan B, Blake GM, et al. Vertebral morphometry: a comparison of long-term precision of morphometric X-ray absorptiometry and morphometric radiography in normal and osteoporotic subjects. OsteoporosInt. 2001;12(2):158–66.

32. Ferrar L, Jiang G, Eastell R. Vertebral wedge angle measured by morphometric X-ray absorptiometry. OsteoporosInt. 2001;12(11):914–21.

33. Szulc P, Munoz F, Marchand F, Delmas PD. Semiquantitative evaluation of prevalent vertebral deformities in men and their relationship with osteoporosis: the MINOS study. OsteoporosInt. 2001;12(4):302–10.

34. Frobin W, Leivseth G, Biggemann M, Brinckmann P. Vertebral height, disc height, posteroanterior displacement and dens-atlas gap in the cervical spine: precision measurement protocol and normal data. ClinBiomech(Bristol, Avon). 2002;17(6):423–31.

35. Shao Z, Rompe G, Schiltenwolf M. Radiographic changes in the lumbar intervertebral discs and lumbar vertebrae with age. Spine. 2002;27(3):263–8.

36. Pfeiffer M, Geisel T. Analysis of a computer-assisted technique for measuring the lumbar spine on radiographs: correlation of two methods. AcadRadiol. 2003;10(3):275–82.

37. Keynan O, Fisher CG, Vaccaro A, Fehlings MG, Oner FC, Dietz J, et al. Radiographic measurement parameters in thoracolumbar fractures: a systematic review and consensus statement of the spine trauma study group. Spine (Phila Pa 1976). 10.1097/01.brs.0000201261.94907.0d doi ;00007632-200603010-00025 pii 2006;31(5):E156–E65.

38. Fink HA, Milavetz DL, Palermo L, Nevitt MC, Cauley JA, Genant HK, et al. What proportion of incident radiographic vertebral deformities is clinically diagnosed and vice versa? JBone MinerRes. 2005;20(7):1216–22.

39. O’Neill TW, Varlow J, Felsenberg D, Johnell O, Weber K, Marchant F, et al. Variation in vertebral height ratios in population studies. European Vertebral Osteoporosis Study Group. Journal of Bone & Mineral Research. Dec 1994;9(12):1895–907.

40. Schwartz EN, Steinberg D. Detection of vertebral fractures. CurrOsteoporosRep. 2005;3(4):126–35.

41. Pflugmacher R, Schroeder RJ, Klostermann CK. Incidence of adjacent vertebral fractures in patients treated with balloon kyphoplasty: two years’ prospective follow-up. Acta Radiol. P22233334L147355 pii ;10.1080/02841850600854928 doi 2006;47(8):830–40.

42. Shindle MK, Gardner MJ, Koob J, Bukata S, Cabin JA, Lane JM. Vertebral height restoration in osteoporotic compression fractures: kyphoplasty balloon tamp is superior to postural correction alone. Osteoporosis International. 2006;17(12):1815–9.

43. Siminoski K, Jiang G, Adachi J, Hanley D, Cline G, Ioannidis G, et al. Accuracy of height loss during prospective monitoring for detection of incident vertebral fractures. Osteoporosis International. 2005;16(4):403–10.

44. Zhao KD, Ben-Abraham EI, Magnuson DJ, Camp JJ, Berglund LJ, An KN, et al. Effect of Off-Axis Fluoroscopy Imaging on Two-Dimensional Kinematics in the Lumbar Spine: A Dynamic In Vitro Validation Study. J Biomech Eng. May 2016;138(5):054502.

45. Zhao KD, Yang C, Zhao C, Stans AA, An KN. Assessment of noninvasive intervertebral motion measurements in the lumbar spine. JBiomechanics. 2005;38(9):1943–6.

46. Reitman CA, Hipp JA, Nguyen L, Esses SI. Changes in segmental intervertebral motion adjacent to cervical arthrodesis: a prospective study. Spine (Phila Pa 1976). Jun 1 2004;29(11):E221–6. Epub 2004/05/29.

47. Reitman CA, Mauro KM, Nguyen L, Ziegler JM, Hipp JA. Intervertebral motion between flexion and extension in asymptomatic individuals. Spine (Phila Pa 1976). Dec 15 2004;29(24):2832–43. Epub 2004/12/16.

48. Patwardhan AG, Havey RM, Wharton ND, Tsitsopoulos PP, Newman P, Carandang G, et al. Asymmetric motion distribution between components of a mobile-core lumbar disc prosthesis: an explanation of unequal wear distribution in explanted CHARITE polyethylene cores. J Bone Joint Surg Am. May 2 2012;94(9):846–54.

49. Pearson AM, Spratt KF, Genuario J, McGough W, Kosman K, Lurie J, et al. Precision of lumbar intervertebral measurements: Does a computer-assisted technique improve reliability? Spine. 2011;36(7):572–80.

50. Hurxthal LM. Measurement of anterior vertebral compressions and biconcave vertebrae. Am J Roentgenol. 1968;103:635–44.

51. Diacinti D, Guglielmi G. Vertebral morphometry. Radiologic Clinics. 2010;48(3):561–75.

52. Genant HK, Wu CY, van KC, Nevitt MC. Vertebral fracture assessment using a semiquantitative technique. JBone MinerRes. 1993;8(9):1137–48.

53. CDC. Examination Staff Procedures Manual for the Health and Nutrition Examination Survey, 1976-1979. Centers for Disease Control and Prevention; 1976.

54. Black DM, Cummings SR, Stone K, Hudes E, Palermo L, Steiger P. A new approach to defining normal vertebral dimensions. J Bone Min Res. 1991;6:883–92.

55. Diacinti D, Pisani D, Del Fiacco R, Francucci CM, Fiore CE, Frediani B, et al. Vertebral morphometry by X-ray absorptiometry: which reference data for vertebral heights? Bone. 2011;49(3):526–36.

56. Djoumessi RMZ, Maalouf G, Wehbe J, Nehme A, Maalouf N, Seeman E. The varying distribution of intra-and inter-vertebral height ratios determines the prevalence of vertebral fractures. Bone. 2004;35(2):348–56.

57. Rea J, Steiger P, Blake G, Potts E, Smith I, Fogelman I. Morphometric X-ray absorptiometry: reference data for vertebral dimensions. Journal of Bone and Mineral Research. 1998;13(3):464–74.

58. Staub BN, Holman PJ, Reitman CA, Hipp JA. Sagittal plane lumbar intervertebral motion during seated flexion-extension in 658 asymptomatic, non-degenerated levels. J Neurosurgery Spine. 2015 2015;23(6):731–8.

59. Fedorov A, Beichel R, Kalpathy-Cramer J, Finet J, Fillion-Robin J-C, Pujol S, et al. 3D Slicer as an image computing platform for the Quantitative Imaging Network. Magnetic resonance imaging. 2012;30(9):1323–41.

60. Shackleford JA, Shusharina N, Verberg J, Warmerdam G, Winey B, Neuner M, et al. Plastimatch 1.6-current capabilities and future directions. MICCAI, First International Workshop on Image-Guidance and Multimodal Dose Planning in Radiation Therapy 2012.

61. Antani S, Cheng J, Long J, Long LR, Thoma GR. Medical validation and CBIR of spine x-ray images over the Internet. Internet Imaging VII: International Society for Optics and Photonics; 2006. p. 60610J.

62. Goh S, Tan C, Price R, Edmondston S, Song S, Davis S, et al. Influence of age and gender on thoracic vertebral body shape and disc degeneration: an MR investigation of 169 cases. The Journal of Anatomy. 2000;197(4):647–57.

63. Cheng X, Sun Y, Boonen S, Nicholson P, Brys P, Dequeker J, et al. Measurements of vertebral shape by radiographic morphometry: sex differences and relationships with vertebral level and lumbar lordosis. Skeletal radiology. 1998;27(7):380–4.

64. McCloskey E, Spector T, Eyres K, Fern E, O’Rourke N, Vasikaran S, et al. The assessment of vertebral deformity: a method for use in population studies and clinical trials. Osteoporosis Int. 1993;3:138–47.

65. Fukunaga M, Nakamura T, Shiraki M, Kuroda T, Ohta H, Hosoi T, et al. Absolute height reduction and percent height ratio of the vertebral body in incident fracture in Japanese women. Journal of bone and mineral metabolism. 2004;22(2):104–10.

66. Lau E, Chan H, Woo J, Lin F, Black D, Nevitt M, et al. Normal ranges for vertebral height ratios and prevalence of vertebral fracture in Hong Kong Chinese: a comparison with American Caucasians. Journal of Bone and Mineral Research. 1996;11(9):1364–8.

67. Eastell R, Cedel SL, Wahner HW, Riggs BL, Melton LJ, III. Classification of vertebral fractures. J Bone Min Res. 1991;6:207–15.

68. O’Brien MF, Kuklo TR, Blanke KM, Lenke LG. Spinal deformity study group radiographic measurement manual. Memphis, TN: Medtronic Sofamor Danek. 2005.

69. Lian J, Levine N, Cho W. A review of lumbosacral transitional vertebrae and associated vertebral numeration. European Spine Journal. 2018;27(5):995–1004.

70. Shah M, Halalmeh DR, Sandio A, Tubbs RS, Moisi MD. Anatomical variations that can lead to spine surgery at the wrong level: Part III lumbosacral spine. Cureus. 2020;12(7).

71. Abola MV, Teplensky JR, Cooperman DR, Bauer JM, Liu RW. Pelvic Incidence in Spines With 4 and 6 Lumbar Vertebrae. Global Spine Journal. 2019;9(7):708–12.

72. Hanhivaara J, Määttä JH, Niinimäki J, Nevalainen MT. Lumbosacral transitional vertebrae are associated with lumbar degeneration: retrospective evaluation of 3855 consecutive abdominal CT scans. European Radiology. 2020;30(6):3409–16.

73. Apazidis A, Ricart PA, Diefenbach CM, Spivak JM. The prevalence of transitional vertebrae in the lumbar spine. The Spine Journal. 2011;11(9):858–62.

74. Bron JL, van Royen BJ, Wuisman P. The clinical significance of lumbosacral transitional anomalies. Acta Orthopaedica Belgica. 2007;73(6):687.

75. Konin G, Walz D. Lumbosacral transitional vertebrae: classification, imaging findings, and clinical relevance. American Journal of Neuroradiology. 2010;31(10):1778–86.

76. Tang M, Yang X-f, Yang S-w, Han P, Ma Y-m, Yu H, et al. Lumbosacral transitional vertebra in a population-based study of 5860 individuals: Prevalence and relationship to low back pain. European journal of radiology. 2014;83(9):1679–82.

77. Nardo L, Alizai H, Virayavanich W, Liu F, Hernandez A, Lynch JA, et al. Lumbosacral transitional vertebrae: association with low back pain. Radiology. 2012;265(2):497–503.

78. Ravi B, Rampersaud R. Clinical magnification error in lateral spinal digital radiographs. Spine. 2008;33(10):E311–E6.

79. Shigematsu H, Koizumi M, Yoneda M, Iida J, Oshima T, Tanaka Y. Magnification error in digital radiographs of the cervical spine against magnetic resonance imaging measurements. Asian spine journal. 2013;7(4):267.

80. Lunt M, Gowin W, Johnell, Armbrecht G, Felsenberg D. A statistical method to minimize magnification errors in serial vertebral radiographs. OsteoporosInt. 2001;12(11):909–13.

